# Selective Decontamination of the Digestive Tract in Invasively Ventilated Patients in an Intensive Care Unit: A protocol for a Systematic Review and Meta-Analysis

**DOI:** 10.1101/2022.03.18.22272586

**Authors:** Naomi E Hammond, John Myburgh, Gian Luca Di Tanna, Tessa Garside, Ruan Vlok, Sajeev Mahendran, Derick Adigbli, Simon Finfer, Fiona Goodman, Gordon Guyatt, Balasubramanian Venkatesh, Ian Seppelt, Anthony Delaney

**Author notes:** **Corresponding author:** Associate Professor Anthony Delaney. **Prospero Registration**: CRD42022309825. Denotes equal senior author (Ian Seppelt, Anthony Delaney).

## Abstract

**Introduction:** The use of Selective Decontamination of the Digestive Tract (SDD) as a preventative infection-control strategy in invasively ventilated patients in the intensive care unit (ICU) remains low despite numerous randomised controlled trials (RCTs) consistently reporting reductions in interval mortality rates and shorter durations of mechanical ventilation. The Selective Decontamination of the Digestive Tract in the Intensive Care Unit (SuDDICU) cluster cross-over RCT, that includes over 5500 participants randomised to receive a standardised commercial grade SDD interventions or standard care, will be reported in 2022 and will add substantive weight to previous RCT data assessing the effect of SDD on interval mortality compared to standard care. We will conduct an updated systematic review and prospective aggregate data meta-analysis of previous conducted and published RCTs, developed using a protocol and statistical analysis plan completed prior to the completion of the SuDDICU RCT and including the SuDDICU data to present the most current evidence available to guide clinical practice.

**Methods and analysis:** We will include RCTs that compare the effect on hospital mortality and other patient-centred outcomes of treatment with SDD compared to standard care in invasively ventilated adults in the ICU. We will perform a search that includes the electronic databases MEDLINE and EMBASE and clinical trial registries. Two reviewers will independently screen titles and abstracts, perform full article reviews and extract study data, with discrepancies resolved by a third reviewer. We will report study characteristics and quantify risk of bias. We will perform random effects Bayesian meta-analyses to provide pooled estimates that SDD improves outcomes, whenever it is feasible to do so. We will evaluate overall certainty of evidence using the Grading of Recommendations Assessment, Development, and Evaluation framework.

**Conclusion:** This updated systematic review and prospective meta-analysis will provide clinicians with an expedited assessment of the totality of current evidence about the effect on mortality of using SDD in mechanically ventilated ICU patients.

## Introduction

Selective Decontamination of the Digestive Tract (SDD) is a preventative antimicrobial infection-control strategy originally described in immunocompromised patients with haematological disorders^1^ and extended to critically ill patients managed in the Intensive Care Unit (ICU) in the 1980s.^2,3^

The principal objective of SDD is to prevent the development of nosocomial pulmonary infections caused by proliferation of potentially pathogenic Gram-negative bacteria and secondary overgrowth with yeasts in the upper gastrointestinal tract, particularly in critically ill patients treated with invasive mechanical ventilation. “Selective” decontamination of the digestive tract is achieved by the application of topical non-absorbable antibiotics, typically an aminoglycoside or peptide antibiotic and an antifungal agent to the oropharynx and gastrointestinal tract combined with a short course of intravenous antibiotics, typically a third-generation cephalosporin or fluoroquinolone.^4^

Whilst systematic reviews of over 40 published randomised controlled trials (RCTs) have consistently reported statistically significant and important reductions in hospital mortality,^5^ duration of mechanical ventilation and hospital-acquired pneumonia,^6-10^ use of SDD remains low. This is due to identified clinician-level barriers to the adoption and implementation of SDD into current clinical practice^11^ and weak recommendations about the role and use of SDD published in multiple iterations of international sepsis guidelines over the last 30 years.^12-14^

Uncertainty about the effectiveness of SDD is due to concerns about the generalisability of RCTs with limited internal and external validity and unsubstantiated concerns about development of antibiotic resistance, particularly to colistin and cephalosporins, with the use of SDD.^15,16^ In addition, there is substantial variation in the concentrations and constitutions of the SDD preparations and duration of application of the intervention.^5^

The Selective Decontamination of the Digestive Tract in the Intensive Care Unit (SuDDICU) randomised controlled trial, conducted in Australia, completed recruitment in November 2021.^17^ This cluster-crossover RCT was conducted in 19 ICUs in Australia and recruited over 5900 patients over two 12-month intervention periods. Apart from the substantive size of the study population, a unique feature of this trial was the use of a commercially manufactured, Good Manufacturing Practice-certified study drug preparation that was applied to all included critically ill ventilated patients for the duration of invasive ventilation during their index ICU admission.^17^

We present the protocol and statistical analysis plan for an updated systematic review and prospective Bayesian aggregate data (trial level) meta-analysis of RCTs that was developed prior to the completion and data analysis of the SuDDICU trial to assess the effect of SDD compared to standard care on interval mortality and other important outcomes in invasively ventilated ICU patients.

## Methods

We will conduct a systematic review of randomised clinical trials with meta-analysis in accordance with the recommendations of the Cochrane Handbook of Systematic Reviews of Interventions^18^ and will report the findings as per the Preferred Reporting Items for Systematic Review and Meta-Analysis (PRISMA) statement.^19^ This systematic review has been registered on the International Prospective Register of Systematic Reviews (PROSPERO Number: CRD42022309825).

### Eligibility Criteria

#### Study types

We will include randomised clinical trials in which the unit of randomisation is individual participants, as well as cluster randomised clinical trials, and cluster cross over clinical trials. There will be no restriction on publication status, language, nor year of publication.

#### Population

We will include trials in which the population includes a sample of critically ill patients of whom ≥75% are invasively ventilated.

#### Intervention

We will include trials in which the intervention is the administration of antibacterial and/or antifungal agents for the duration of mechanical ventilation administered via the oral, nasal, or gastric route to the upper gastrointestinal tract, stomach or proximal small bowel, with or without the administration of systemic antibiotics.

#### Comparison

We will include trials in which the comparison group is either allocated to placebo or standard care.

#### Outcomes

We will include studies that report any of the outcomes specified for this review.

#### Exclusion criteria

- Trials that solely use oral antiseptic agents
- Trials that will not have data available for inclusion in a timely fashion

### Information sources

We will perform a search of the electronic databases MEDLINE and PreMEDLINE, EMBASE, CinAHL and the Cochrane Central Register of Clinical Trials to identify published trials and conference abstracts. We will search clinical trial registries through the World Health Organisation international clinical trials registry platform. We will search for unpublished trials by searching the PubMed indexed pre-print servers, as well as through contacting experts in the field. We will manually search reference lists of included studies and other systematic reviews.

### Search strategy

We will develop our search strategy in alignment with the Peer Review of Electronic Search Strategies (PRESS) guideline statement.^20^ We will use a combination of search terms to identify critically ill patients, mechanical ventilation and Selective Digestive Decontamination (SDD)/Selective Oral Decontamination (SOD) and combine them with sensitive filters to identify randomised clinical trials^18^ including cluster and cross over trials.

### Data management and selection process

Study records identified by the search will be downloaded into a reference management system software.^21^ Duplicate records will be removed. Two authors will independently screen the titles and abstracts of identified study records to identify potentially eligible studies. Differences will be resolved by discussion or by a third reviewer following which we will retrieve full text manuscripts of studies that are deemed potentially eligible for detailed review.

### Selection process

Two authors will independently apply the eligibility criteria to all potentially eligible studies. Differences will be resolved by discussion or by a third reviewer. Reasons for exclusion will be recorded and presented in a Preferred Reporting Items for Systematic Reviews and Meta-analyses (PRISMA) flow diagram.^19^

### Data collection process

Data extraction forms will be developed, and pilot tested in consultation with the statisticians. Data will be extracted from the included studies onto specific data collection forms. Data extraction will be performed in duplicate, with discrepancies resolved by discussion or by a third reviewer. If required, we will contact authors for further essential information, and cross check data against existing secondary sources.

### Data items

We will extract data regarding study characteristics (including first author, year of publication, unit of randomisation, study type, number of participants, number of sites), details of the included ICUs (e.g., medical, surgical, trauma, mixed general ICU) and participants (including age distribution, sex distribution, duration of ventilation at baseline, severity of illness), details of the interventions (topical agents, dose and duration; systemic agents, dose and duration) as well as details of the comparison group (placebo or usual care, use of topical antiseptics, use of VAP bundles).

### Risk-of-Bias assessment

Two review authors, with no affiliation with any of the included RCTs, will independently assess the risk of bias for each included study. To assess risk of bias, we will use all publicly available reports of trials, including published trial protocols and statistical analysis plans. Disagreements will be resolved by discussion, involving a third independent assessor if needed. We will use the McMaster University CLARITY Group ‘Tool to Assess Risk of Bias in Randomized Controlled Trials’ with modifications to account for cluster and cross-over trials.^22^ Details regarding the risk of bias assessments will be reported in the electronic supplement.

### Outcomes

#### Primary

The primary outcome will be hospital mortality (or closest approximation provided)

#### Secondary

The Secondary outcomes will be:

- Duration of mechanical ventilation
- Incidence of antibiotic resistant micro-organisms reported in the ICU (as defined and reported by each trial)
- Incidence of antibiotic resistant micro-organisms in patients (as defined and reported by each trial)
- Incidence of ventilator associated pneumonia (as defined and reported by each trial)
- Incidence of Clostridioides difficile in patients and in units/clusters/wards (as defined and reported by each trial)
- ICU length of stay
- Hospital length of stay
- Mortality at the longest time point reported

### Data analyses

The main analyses will be performed on all included trials regardless of risk of bias. For binary outcomes, we will use risk ratios (RR) while for continuous outcomes mean differences (MD) or standardised MD. We will perform random effects Bayesian meta-analyses to provide pooled estimates. Along with the pooled effect sizes and 95% credible intervals, we will report the posterior probabilities that SDD is associated with improved outcomes compared to standard care (for each outcome analysed). We will perform sensitivity analyses examining treatment effects using different priors including vague and weakly-informative priors on effect and heterogeneity parameters. As a vague prior for the mean effect we will use the unit information prior, that is, for the log RR a normal distribution with mean 0 and standard deviation 2: this corresponds to RRs within a range of 1/50 to 50 with 95% probability. For the heterogeneity parameter we will use a half-normal (0.5) distribution as a vague prior and the appropriate weakly informative log-normal heterogeneity prior for binary outcomes.^23,24^

As an additional sensitivity analysis, we will perform frequentist random-effects meta-analysis by using Hartung-Knapp-Sidik-Jonkman^25^ and Der-Simonian Laird estimates of the between-study variance.

As some of the included trials are cluster randomized trials we will take account of clustering by adjusting the raw data for the design effect by using the effective sample size approach (i.e. the original sample size is divided by the design effect which is 1+(average cluster size-1)*Intracluster Correlation Coefficient.^26^ Analyses will be based on reported data and we will try to obtain missing outcome data from the original study authors. Complete case analyses will be performed as we will not impute any missing data.

### Assessment of heterogeneity

We will assess quantitative heterogeneity by reporting the posterior estimates of the heterogeneity parameter (tau^2^) with its 95% credible interval, the prediction interval of the intervention pooled effect size and evaluating the proportion of total variability due to heterogeneity rather than due to sampling error (I^2^).

### Assessment for small study/publication bias

We will evaluate small-study effects by visual assessment of the contour-enhanced funnel plots and formal Egger’s regression test.

All pooled results will be presented in the form of forest plots. All statistical analyses will be performed using R (for the Bayesian meta-analysis using the package bayesmeta^27^) and Stata.

### Predefined subgroup analysis

The primary outcome hypothesis is that SDD will reduce mortality compared with standard care/placebo in invasively ventilated adult patients.

We will restrict subgroup analyses to the primary outcome including the following:

- SDD with oral and/or enteral agents only v SDD that includes oral, enteral, and intravenous agents as the intervention. We hypothesize that any reduction in mortality will be greater with use of intravenous agents compared to oral and/or enteral agents alone.
- Trials conducted in medical ICUs compared to Surgical ICUs compared to trauma ICUs compared to mixed population/ICUs. We hypothesize that the any reduction in mortality will be greater in surgical patients compared to medical or mixed populations.
- Individual patient vs unit level randomisation (i.e. cluster and cluster/cluster-cross-over). The two hypothesis for the direction of this subgroup effect are 1) any reduction in mortality associated with the use of SDD compared to placebo will be greater in individual patient randomised trials compared to unit level randomised trials, and 2) Any increase in antibiotic resistant organisms will be greater in unit level randomisation compared to individual patient randomisation.

The credibility of any subgroup analysis will be assessed using the Instrument to assess the Credibility of Effect Modification Analyses (ICEMAN) in randomized controlled trials and meta-analyses.^28^

## Confidence in cumulative evidence

We will use the Grading of Recommendations Assessment, Development, and Evaluation (GRADE)^29^ approach to assess the overall certainty of evidence that SDD compared with placebo or standard care improves outcome for each primary and secondary outcome measure and present the results in a ‘summary of findings’ table^30^ including plain language summary.^31^ The certainty of evidence will be evaluated by the discussion with all authors and be based on the methods in the Cochrane Handbook of Systematic Reviews and updated GRADE working group recommendations.^29,32,33^ The overall certainty of evidence for each outcome will be rated “high”, “moderate”, “low” or “very low”. We will develop summary of finding tables using optimal formats,^30^ presenting both relative and absolute effects and including plain language summaries with wording following GRADE guidance.^31^

## Data Availability

All data produced in the present work are contained in the manuscript

## Ethics and dissemination

This review does not require ethical approval as this is a systematic review of published studies. We plan to present the results of the systematic review at national and international scientific meetings and will prepare a manuscript for submission to a peer reviewed journal.

## Discussion and limitations

This systematic review and meta-analysis will provide the most up-to-date synthesis of evidence on the effect of the use of topical and/or enteral antibiotics or antifungals compared with placebo or usual care on hospital mortality and other secondary and exploratory outcomes in patients invasively ventilated in an Intensive Care Unit.

We acknowledge that there will be limitations to the proposed systematic review, including that eligible studies are anticipated to be heterogeneous in nature due to variations in the interventions given, trial design, and variation in treatment of the comparison groups. In addition, the strength of a systematic review and meta-analysis relies in part on the strength of included studies, and therefore may be limited due to the paucity of high quality randomised controlled trials in this area.

